# Developing an intervention prototype to support people with multimorbidity in addressing social care needs: What factors should be considered?

**DOI:** 10.1101/2025.10.08.25336767

**Authors:** Sian Holt, Lucy Smith, Miriam Santer, Firoza Davies, Andrew Farmer, Glenn Simpson, Chika Nwokedi, Hajira Dambha-Miller, Leanne Morrison

## Abstract

**Background:** People living with multimorbidity often face complex social care needs that significantly affect their health and wellbeing. Despite growing recognition of the importance of addressing these needs in primary care, practical and systemic barriers, such as time constraints, unclear professional roles, and fragmented service pathways, limit effective support.

**Methods:** Using the Person-Based Approach, we developed a dual-component intervention: (1) a brief screening tool for primary care professionals (PCPs) to identify patients with potential social care needs, and (2) a patient-facing self-navigation tool to help individuals recognise, prioritise, and plan responses to their needs. Sixteen patients and fifteen PCPs participated in think-aloud interviews. Data were analysed using a Table of Changes to inform real-time optimisation.

**Results:** Participants appreciated the autonomy-supportive, personalised tone of the self-navigation tool and its journey-based framing. Managing expectations was essential to avoid misinterpretation, and acknowledging prior negative healthcare experiences helped build trust. PCPs valued the screening tool’s brevity and practicality but raised concerns about role clarity and integration into workflows. Both groups highlighted the importance of accessibility, digital inclusion, and the burden of multimorbidity.

**Conclusion:** Findings informed a practical checklist for future intervention design, including: (1) prioritising control and choice, (2) setting clear expectations, (3) minimising burden, (4) acknowledging past experiences, (5) clarifying roles, and (6) embedding tools within existing systems. This study offers actionable insights into person-centred intervention development and highlights the need for further evaluation to assess effectiveness and scalability in improving outcomes and system efficiency.

## Introduction

In the UK, over a quarter of the population are living with multimorbidity (1). The number of people living with multimorbidity is expected to rise in the future, with more young and middle-aged adults affected (2, 3). People living with multimorbidity account for 53% of GP consultations in the UK (1). The management of multimorbidity is more complex than for single conditions, requiring input from various health and social care services that can challenge continuity of care (4, 5). People living with multimorbidity may struggle to prioritise care for their conditions and needs (6), whilst limited resources may undermine timely multidisciplinary working among professionals (7). Systematic review evidence shows that multimorbidity has a large economic burden worldwide (8). The World Health Organisation has highlighted the role of primary care in managing multimorbidity (9).

People living with multimorbidity frequently encounter a wide range of social care needs that can complicate the management of their health and wellbeing. These needs extend beyond clinical care and encompass issues such as secure and adequate housing, employment instability, financial insecurity, limited mobility, difficulties with activities of daily living (e.g., shopping, personal hygiene, self-care), and reduced access to community or social support networks (10-13). Social isolation and reduced engagement with meaningful social roles are also common, particularly as health conditions accumulate (14, 15). The presence of unmet social needs in multimorbidity has been linked to poorer health outcomes, including diminished quality of life, increased rates of hospitalisation, and higher mortality (16). Furthermore, when social needs remain unaddressed, they can contribute to a greater burden on healthcare services and increased costs for both the NHS and wider society, due to more frequent healthcare utilisation and potentially preventable admissions (17).

Despite increasing recognition of the broader determinants of health, addressing social and contextual needs in primary care remains challenging. Barriers include time constraints, limited resources, and the complexity of navigating health and social care systems, which hinder systematic identification and support for patients with SCNs during routine consultations (18). Population clustering approaches have been demonstrated as an efficient method for identifying individuals at risk of SCNs among those with multimorbidity (19). Interventions are now required to integrate these clustering methods into primary care workflows. A recent qualitative scoping study explored perspectives of individuals with multimorbidity, carers and healthcare professionals regarding the application of AI-derived clusters to improve SCN-related care in primary care settings (20). Findings indicated that the approach is considered valuable when integrated with effective clinical communication and personalised care. Previous research also emphasises the importance of healthcare professionals acknowledging, discussing, and supporting patients in relation to their SCNs (13, 21). The present study aimed to identify key behavioural and contextual factors that can influence engagement with interventions designed to address social care needs in primary care settings.

## Methods

As part of a broader research programme (22), a prototype intervention was developed to facilitate the identification and management of social care needs in primary care for individuals with multimorbidity. Development followed the Person-Based Approach, with emphasis on acceptability, feasibility, and engagement (22). The prototype intervention comprised two complementary components: (1) a primary care-facing tool incorporating an artificial intelligence algorithm to identify patients at increased risk of unmet social care needs, and (2) a patient-facing self-navigation tool to support recognition, prioritisation, and planning of responses to social care needs. The components were designed to enable brief, targeted discussions on social care needs during routine primary care consultations.

### Design

We conducted semi-structured qualitative think-aloud interviews to identify key behavioural and contextual factors influencing engagement with this intervention at the prototype stage. Interviews took place between February and June 2025.

### Participants

Two groups were recruited: adults (≥18 years) with multimorbidity, defined as ≥2 physical and/or mental health conditions from a stakeholder-validated list of 59 (23), and primary care professionals. Individuals lacking the capacity to give informed consent (e.g. cognitive impairment) were excluded from the study.

Participants with multimorbidity were recruited via research-active general practices using posters in waiting rooms and patient newsletters, supplemented by social media outreach and the Voice online platform (24). Primary care professionals were recruited through the Regional Research Delivery Network, elevator pitches at PCP meetings, and LinkedIn advertisements. Interested individuals accessed the study website to register or contacted the lead researcher. Recruitment closed upon reaching the target sample size, which was determined based on previous research. Data saturation was reached when no substantial new information was identified in the final interviews (25, 26).

### Procedure

People living with multimorbidity and PCPs who were interested in taking part accessed the study website (hosted on Qualtrics), which included the participant information sheet, an online consent form and demographic survey (27). Information was collected including sex, gender, ethnicity, education, occupation, and types of long-term conditions. Responders were purposively sampled across these factors to ensure a diverse range of views were included.

Selected participants were invited to an online or telephone interview at a time and date of their choosing. Translation services were available for participants who wished to be interviewed in a language other than English; however, no participants chose this. Each participant was given a £15 shopping voucher to thank them for their time.

Interviews ranged from 41 minutes to 65 minutes for people living with multimorbidity and 19 minutes to 53 minutes for PCPs. Interviews were conducted by an experienced qualitative researcher (SH). SH is a White British female postdoctoral researcher in her 30s with lived experience as a carer to her husband who lives with multimorbidity. These factors may influence how the interviews were conducted and analysed, including a more empathetic interview style and sensitivity to disclosures relevant to lived experiences. Interviews were audio recorded and transcribed verbatim.

Topic guides were developed with input from our co-authors and Lived Experience Advisory Panel (LEAP), and piloted prior to use with PPI members and GP trainees. The topic guide opened with an exploratory question about current experiences with seeking help for social care needs (patients) or discussing social care needs (PCPs). A think-aloud of the intervention components (social care needs identification tool and patient self-navigation tool) followed. Participants were asked to read intervention content out loud and verbalise their thoughts, comments and reactions. Neutral prompts encouraged elaboration on reactions to explore the behavioural and contextual factors influencing engagement. The prototypes were refined iteratively in parallel to interviews. All changes made were documented in a Table of Changes (28) and prioritised in collaboration with stakeholders and the LEAP group.

### Analysis

The primary analysis of the data used the Table of Changes to prioritise iterative changes to the intervention prototype. Throughout this process, we identified key behavioural and contextual factors influencing engagement with the intervention.

### Patient and Public Involvement

Monthly meetings were held with a Lived Experience Advisory Panel (LEAP) comprising two members with experience of multimorbidity, neurodiversity, and cultural diversity. Discussions addressed patient-facing materials, study protocol, recruitment strategies, community engagement, intervention development, data analysis, and dissemination. One LEAP member, also a core PPI contributor, participated in wider research team meetings to provide ongoing input. A primary care professional advisory group was also convened, consisting of a general practitioner, social worker, community well-being services manager, and well-being coach, all external to the core research team. This group provided iterative feedback on recruitment strategies and intervention prototypes.

A Patient and Public Involvement and Engagement (PPIE) event was held on 6 June 2025 to elicit feedback on the patient-facing self-navigation component of the intervention. The event design incorporated measures to address participant requirements. Feedback obtained in this setting provided additional qualitative data from a diverse participant group, which was subsequently incorporated into iterative content refinement. Attendees were five people either living with multimorbidity or caring for someone with multimorbidity. Demographics were not recorded for this group.

## Results

### Sample characteristics

We interviewed 16 people living with multimorbidity (Table 1). Most participants were female (n=10) and half were educated to a postgraduate level (n=8). A range of age groups participated, and a selection of ethnic groups were represented (e.g. Caribbean, African, Pakistani and Mixed). People living with multimorbidity reported an average of four conditions (ranging from two to nine total conditions). The most common conditions were depression/anxiety, chronic primary pain, osteoporosis/osteoarthritis, post-traumatic stress disorder, diabetes and stroke.

**Table 1:** Demographics for people living with multimorbidity.

| **Demographics** | **(n=16)** | **%** |
| --- | --- | --- |
| **Sex** |  |  |
| Female | 10 | 63% |
| Male | 5 | 31% |
| Prefer not to say | 1 | 6% |
| **Age** |  |  |
| 18 to 29 | 1 | 6% |
| 30 to 39 | 4 | 25% |
| 40 to 49 | 4 | 25% |
| 50 to 59 | 5 | 31% |
| 60 to 69 | 2 | 13% |
| **Ethnicity** |  |  |
| White British | 10 | 63% |
| White – Any other White background | 1 | 6% |
| Black or Black British - Caribbean | 1 | 6% |
| Black or Black British - African | 1 | 6% |
| Asian or Asian British - Pakistani | 2 | 13% |
| Mixed – Any other mixed background | 1 | 6% |
| **Education** |  |  |
| GCSE | 3 | 19% |
| A Levels | 2 | 13% |
| Undergraduate | 3 | 19% |
| Postgraduate | 8 | 49% |

We also interviewed 15 primary care professionals (Table 2). Participants were GPs (n=7), practice nurses (n=3), social prescribers (n=2), health/wellbeing coaches (n=1), health and wellbeing team leads (n=1) and care coordinators (n=1). Most participants were female (n=12) and White British (n=13), with some representation of Indian ethnicity (n=2).

**Table 2:**
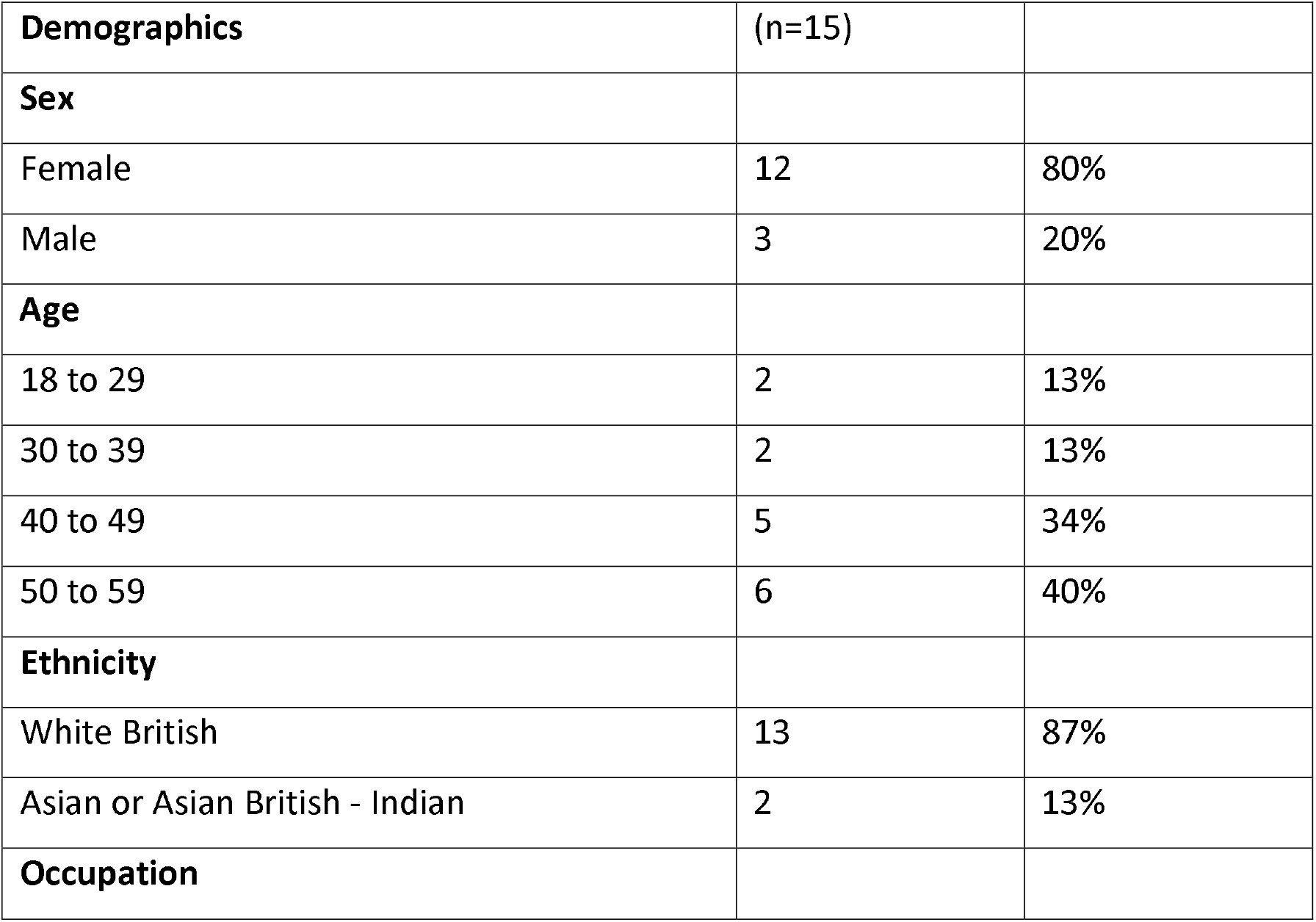
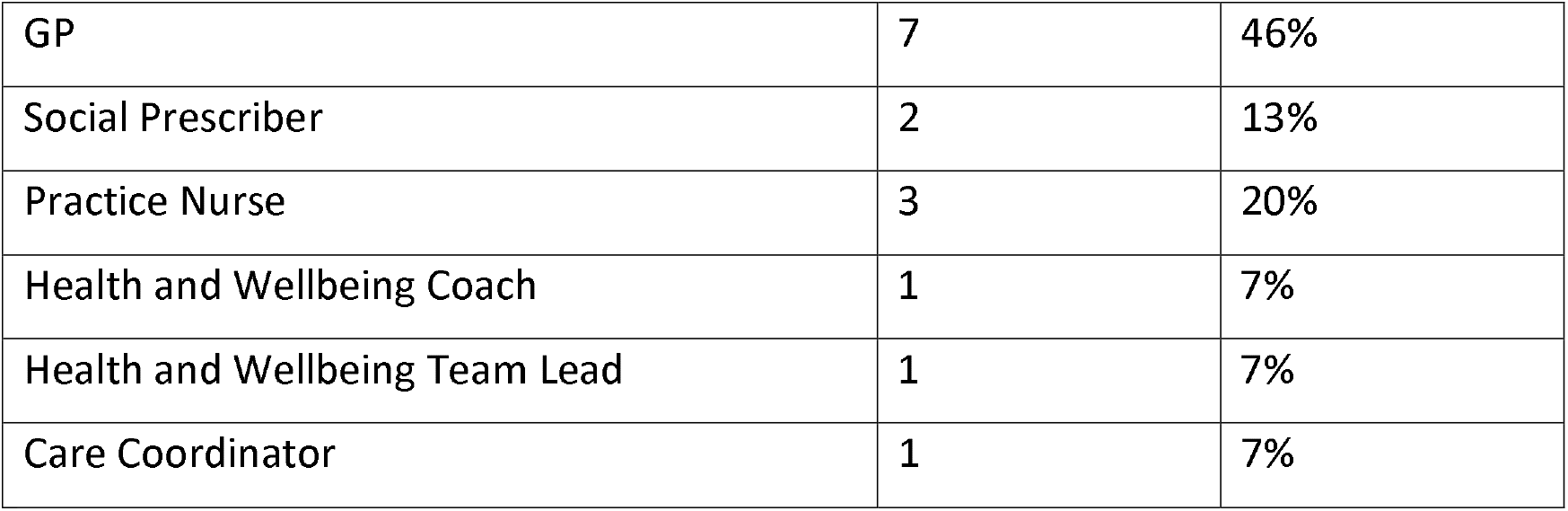
Demographics for primary care professionals.

### Behavioural and contextual factors shaping engagement

Analysis of think-aloud interviews with people living with multimorbidity and primary care professionals (PCPs), in addition to PPIE event attendees, identified 6 behavioural and contextual factors that shaped engagement with the intervention prototype.

#### 1. Maintaining control and choice

People living with multimorbidity appreciated the supportive and autonomy-focused language used in the self-navigation tool, offering the choice between recommendations (made by the AI-clustering algorithm) or exploring their own priorities at their own pace. The ability to select motivations and priorities fostered a sense of ownership and control. It also made the intervention feel more personal and specific to their own experiences, which they felt could be more useful than generalized approaches.

> *“I like the word ‘my’, so it gives you ownership… it’s giving you autonomy*.*”* (MLTC-10)
>
> *“I like the sound of that*… *Because it’s personal to you. It’s how you feel and what would work for you, rather than it’s a generic thing that is supposed to be across the board*.*” (MLTC-16)*

People living with multimorbidity had an overall positive response to the caring tone and personal language used, valuing the intervention treating them as an individual. The language made this intervention feel different to other interventions they had experienced previously, being perceived as potentially more useful. This was particularly important for individuals who had previously felt disempowered in healthcare settings.

> *“It sounds like it’s actually going to be something that’s going to be helpful, and it sounds caring, it doesn’t sound just like bog-standard information. It sounds like someone has put thought behind it*.*”* (MLTC-03)

#### 2. Expectation management

A recurring finding was the need to clearly communicate what the self-navigation tool is and what it is not. Several people living with multimorbidity initially assumed it would be a directory of services, leading to confusion and disappointment. Clarifying that the tool is designed to support planning and navigation, rather than providing static service listings, was essential. This highlights the importance of setting expectations early to avoid disengagement and build trust.

> *“I want to see who I contact and why. That’s where I was thinking it was going to go. I don’t know*.*”* (MLTC-11)
>
> *“It would be helpful to say upfront that this isn’t a list of services, but something to help you think through what you need and how to find it*.*”* (MLTC-02)

As a result, clarifications were added to manage expectations of the role of the self-navigation tool as a resource for planning and navigating care, as opposed to listing services which may rapidly become outdated.

Expectation management was also key for the Primary Care Professionals. Initially, the identification tool was framed as a conversation tool. However, PCPs associated the word ‘conversation’ with a fear of ‘opening a can of worms’. They felt that having these conversations would be difficult to control within time constraints of routine appointments.

> *“What if you uncover something? What if you open up a massive can of worms and the patient reveals worrying things*… *often doctors do shrink back from the thought of uncovering something else*.*”* ***PCP-10***

To manage the expectations of primary care professionals we instead framed this as a screening tool. This made the tool comparable to existing processes that are simple, brief and easy to use, making the inclusion of social care needs within routine consultations seem achievable.

#### 3. Burden and overwhelm associated with multimorbidity

People living with multimorbidity felt that improvements to the self-navigation tool could improve the flow and minimize the burden of engaging with the intervention. People living with multimorbidity often felt overwhelmed with the need to manage their own complex health conditions alongside their social care needs. They appreciated the friendly framing of this as an individual journey, which reduced perceptions of pressure and overwhelm.

> *“It’s not lecturing you, it’s not putting too much stress on you, it’s not putting timelines like, I should be further than I am… sometimes you do compare with people, but this is like an individual journey*.*” (MLTC-10)*

Both people living with multimorbidity and primary care professionals raised several accessibility concerns that they felt could impact the flow of navigation and add to cognitive load and burden experienced. They suggested the use of diverse imagery, clear language for non-native English speakers, compatibility with screen readers, use of colors and adding video content for those with different learning styles to increase engagement. They would also prefer the intervention to be adaptable across both digital and non-digital formats for easier access e.g. app, website and leaflet. Most primary care professionals highlighted the need for support to be accessible, with some patients not able to use digital formats.

#### 4. Trust and past experiences

People living with multimorbidity expressed general frustration with the over-stretched healthcare system, perceiving a transferred burden of the need to self-manage their social care needs.

> *“It slightly frustrates me, angers me that this resource has to be created because the system itself is so challenging to navigate*… *that really your GP or somebody else should really be making the plan instead of putting that burden on the patient*.*”* ***MLTC-12***

As part of iterative content changes, we created a section providing problem solving strategies (“I have tried to get help before”). People living with multimorbidity felt validated by this section of the intervention, addressing common challenges like not feeling listened to or difficulties in accessing services.

> *“I like how it says things don’t always go to plan. That’s true*… *I really like that*.*”* ***MLTC-10***

*Acknowledgement* of these previous negative experiences was important for people living with multimorbidity and helped motivate them to continue to engage with the intervention.

#### 5. Perceived relevance and roles

PCPs recognised the importance of social care needs and the impact this has on health for patients.

> *“I’m really pleased that it’s been acknowledged about the need for social care within primary care*… *this research is really important*.*”*⍰***PCP-13***

However, there was a lack of consensus around which PCP should use the tool.

> *“Are they [GPs] going to remember another thing? I don’t know. But would a specially-trained social prescriber be able to do this well? Yes, I think so*.*”* ***PCP-03***
>
> *“it’s good that it’s including GPs as well, because they can equally have the conversation with patients and maybe not just referring on to the social prescriber… from a wider perspective, it’s a good idea to be accessing GPs, nurses, healthcare assistants”* ***PCP-05***

Many PCPs felt frustrated in addressing social care needs due to systematic limitations including a lack of time in routine consultations or available resources/services going ‘out of date’ (e.g. closing or containing information that is no longer relevant). Primary care professionals state that ‘*you know it’s an issue, but you feel a bit powerless to do much about it”* (PCP-07). GPs felt that their training was focused on medical issues, and they might not be able to properly support social care needs in the same way.

> *“[Social care needs] are often very complex. We deal with a lot of complexity in primary care, but it’s medical complexity predominantly, that’s what my training is*.*”* ***PCP-07***

PCPs welcomed the idea of a brief tool that could be a simple and non-burdensome way to identify and support patients with social care needs in primary care.

> *“I think the screening tool is helpful because it’s literally*… *it’s two questions. Really short and very easy to access and it does take away that burden*.*”* ***PCP-05***

They also appreciated the patient-facing self-navigation resource, which they felt could help patients understand and communicate their own needs, which in turn could improve consultations.

#### 6. Embedding within existing systems

Some PCPs were wary of adding additional pop-ups into patient notes systems to help identify social care needs, feeling it may distract from the consultation. Embedding the social care need identification component of the intervention into practice would need careful development to ensure a good fit within the consultation and existing systems in use, in order to facilitate PCP engagement.

> *“I would say that we get so many things popping up that we sometimes get a bit of click fatigue and just end up clicking through things without really properly taking them in*.*” (PCP-08)*
>
> *“I think, the trouble with pop-ups is they can distract from conversations. I’ve had it before… Suddenly the consultation agenda has changed from being the patient’s, to yours, so I do worry about some of these things sometimes… you have to be careful not to interrupt the consultation flow*.*” (PCP-01)*

## Discussion

This study aimed to explore behavioural and contextual factors influencing engagement with a prototype intervention to support the identification and management of social care needs in primary care for individuals with multimorbidity. Maintaining control and choice, expectation management, burden and overwhelm associated with multimorbidity, trust and prior experiences, perceived relevance and professional roles, and integration within existing systems were identified as important factors influencing engagement with the identification and management of social care needs. These findings informed iterative refinements to the intervention which enabled changes to be experienced and optimised in real time. This provides transferable insights for the design and optimisation of data-driven approaches to addressing social care needs in primary care.

People living with multimorbidity responded positively to the tone used in the self-navigation tool, reflecting their values of control and choice. This resonates with the Person-Centred Nursing Framework (29), which emphasizes the importance of respecting individual values. This also reflects the principles of the Person-Based Approach, grounded in self-determination theory, which suggest that intrinsic motivation to engage with interventions is improved by a need for autonomy (30). In this study, people living with multimorbidity valued the choice to explore either AI-derived recommendations or self-selected priorities. This reflects a shift towards facilitating people living with multimorbidity to become active agents in their care which is a key tenet of self-management in chronic illness models (31).

Some people living with multimorbidity misunderstood the tool’s purpose, expecting a directory of services. This highlighted the importance of clear expectation-setting and reinforced the need for tools to support planning and navigation, going beyond static signposting that can rapidly become outdated (32). Managing expectations was also important for primary care professionals. Framing of the intervention by naming it similarly to already known tools (e.g. screening tools) was essential in ensuring primary care professionals felt this would fit within their existing routine consultations.

This study highlighted the careful balance needed when developing interventions for people living with multimorbidity, particularly when they are asked to engage in complex decision making alongside management of their health conditions (33). Friendly framing of this helped to reduce barriers associated with feeling under pressure or overwhelmed. This was also highlighted in discussions around accessibility. Patient and PCP emphasis on accessibility such as plain language, diverse imagery, and multi-format availability, reflects broader concerns about digital exclusion and health disparities (34). These considerations are consistent with findings from community-based interventions that stress the importance of culturally and linguistically appropriate materials to ensure equitable engagement (35).

Participants valued the problem-solving sections, particularly where the tool acknowledged previous negative experiences with the healthcare system and offered solutions, increasing patient self-efficacy in their ability to think-through their own social care needs. This aligns with Bandura’s theory of self-efficacy, which posits that individuals are more likely to engage in health-promoting behaviours when they feel capable of overcoming barriers (31). By validating lived experiences and offering actionable strategies, the self-navigation tool enhanced users’ confidence in their ability to navigate complex social and healthcare systems, which is a critical factor in the management of multimorbidity.

PCPs recognised the importance of social care needs but described significant barriers to addressing them in practice, including time constraints and limited knowledge of local services. Furthermore, there was no clear consensus on which professional group should use the tool, and concerns were raised about the risk of embedding the tool and the potential disruptive impact on consultation flow. This echoes broader implementation science literature, which emphasises the importance of aligning new interventions with existing workflows, clarifying roles, and ensuring that tools are adaptable to local contexts (36). The lack of consensus on who should administer the tool also reflects ongoing debates about safe task-shifting and interprofessional collaboration in primary care settings (37). These findings underscore the need for careful integration of digital tools into existing systems and workflows within primary care.

### Strengths and Limitations

A key strength of this study is the use of the person-based approach, which enabled in-depth exploration of user experiences and informed real-time optimisation of the intervention. The inclusion of both people living with multimobidity, a range of primary care professionals and PPIE community members ensured that diverse perspectives were captured. Ongoing engagement with a lived experience advisory panel and primary care professional advisory group further strengthened the relevance and rigour of the findings.

However, the study has limitations. Participants were self-selecting and may have been more motivated or digitally literate than the broader population. The think-aloud method, while rich in insight, may not fully capture how users engage with tools in real-world settings. Additionally, the intervention was tested in prototype form; further evaluation is needed to assess its impact when implemented at scale.

### Implications for Research and Practice

We present an actionable checklist for future researchers to consider when developing social care needs interventions for patients with multimorbidity in the primary care context.

1. **Prioritise language that reflects maintaining control and choice** to validate individual experiences and autonomy.
2. **Set clear expectations early** using familiar terminology to clearly communicate the purpose.
3. **Minimise burden and overwhelm of multimorbidity** by non-judgmental framing, simple navigation and inclusive distribution (using digital and non-digital methods).
4. **Acknowledge past experiences and offer solutions** to build trust and facilitate user self-efficacy.
5. **Clarify roles and responsibilities considering** flexibility in delivery and training opportunities.
6. **Embed within existing systems** ensuring tools support the flow of routine consultations, co-developing integration strategies with end-users to refine fit and usability.

Further work is needed to develop and evaluate the effectiveness of similar tools in improving social care need identification, patient outcomes, and system-level efficiency.

## Conclusion

This study identified key behavioural and contextual factors influencing engagement with an intervention designed to address social care needs in primary care for people living with multimobidity. Through a person-based, iterative development process, we found that people living with multimorbidity valued personalised, autonomy-supportive tools that acknowledged their lived experiences, while primary care professionals emphasised the need for brevity, role clarity, and seamless integration into existing workflows. As health systems increasingly seek to address the broader determinants of health, our findings provide an actionable checklist for future intervention development for embedding social care need support into routine primary care.

## List of abbreviations

PCPs: primary care professionals
NHS: National Health Service
NIHR: National Institute of Health Research
GP: General Practitioner
PBA: Person-based Approach
LEAP: Lived Experience Advisory Panel

## Acknowledgements

We would like to thank all participants for sharing their thoughts on the intervention prototype. We would also like to thank the South-Central Research Delivery Network for their recruitment support. Finally, we would like to thank our LEAP group contributors and primary care professional advisory group for their input throughout the study.

## Author contributions

LM and HDM were principal investigators for the study and conceptualized the study idea and design. SH and LM designed the intervention prototype with input from all co-authors, LEAP group and primary care professional advisory group. SH was responsible for conducting the study including data collection, analysis and prototype iteration. LM, LS and MS supported Table of Changes analysis. AF, MS, and GS provided wider study support and insights into application of findings. FD (PPI) contributed to LEAP group meetings, including supporting recruitment and discussing findings in relation to lived experiences. SH led the writing of the manuscript. All authors contributed to the final version of the manuscript. All authors read and approved the final manuscript.

## Statements and declarations

### Ethical considerations

Ethical approval was granted by the Research Integrity and Governance team and Faculty of Medicine Ethics Committee at the University of Southampton (99871/100542) and IRAS/HRA (347934).

### Consent to participate

Written consent obtained.

### Consent for publication

Written consent obtained.

### Data availability statemet

Data are available upon reasonable request.

